# Modeling Tempo of COVID-19 Pandemic in India and Significance of Lockdown

**DOI:** 10.1101/2020.05.15.20103325

**Authors:** Brijesh P. Singh, Gunjan Singh

**Affiliations:** Department of Statistics, Institute of Science, Banaras Hindu University, Varanasi-221005; School of Applied Sciences, Amity University, Lucknow-226028

## Abstract

A very special type of pneumonic disease that generated the COVID-19 pandemic was first identified in Wuhan, China in December 2019 and is spreading all over the world. The ongoing outbreak presents a challenge for data scientists to model COVID-19, when the epidemiological characteristics of the COVID-19 are yet to be fully explained. The uncertainty around the COVID-19 with no vaccine and effective medicine available until today create additional pressure on the epidemiologists and policy makers. In such a crucial situation, it is very important to predict infected cases to support prevention of the disease and aid in the preparation of healthcare service. In this paper, we have tried to understand the spreading capability of COVID-19 in India taking into account of the lockdown period. The numbers of confirmed cases are increased in India and states in the past few weeks. A differential equation based simple model has been used to understand the pattern of COVID-19 in India and some states. Our findings suggest that the physical distancing and lockdown strategies implemented in India are successfully reducing the spread and that the tempo of pandemic growth has slowed in recent days.

## Introduction

The Novel Corona Virus (COVID-19) started from Wuhan, China and thus, initially known as the Wuhan virus, expanded its circle in South Korea, Japan, Italy, Iran, USA, France, Spain and finally spreading in India. It is named as novel because it is never seen before mutation of animal corona virus but certain source of this pandemic is still unidentified. It is said that the virus might be connected with a wet market (with seafood and live animals) from Wuhan that was not complying with health and safety rules and regulations. The pandemic COVID-19 have been recorded over 200 countries, territories, and areas with about 3000000 confirmed cased and 200000 deaths (WHO).

The COVID-19 is very similar in symptomatology to other viral respiratory infections. As it is novel virus, the specific modes of transmission are not clearly known. Originally it is emerged from animal source then spread all over the world from person to person. Initially, there has been speculation about the virus spreading while the carrier (infected person) is not showing any symptoms, but that has not been confirmed as a scientific fact (Kachroo, 2020). On 11 March 2020, WHO changed the status of the COVID-19 emergency from public health international emergency to a pandemic? Nonetheless, the fatality rate of the current pandemic is on the rise (between 2–4 percent); relatively lower than the previous SARS-CoV (2002/2003) and MERSCoV (2012) outbreaks (Malik et al., 2020). Thus, COVID-19 has presented an unprecedented challenge before the entire world.

Symptoms of COVID-19are reported as cough, acute onset of fever and difficulty in breathing. Out of all the cases that have been confirmed, up to 20% have been deemed to be severe. Cases vary from mild forms to severe ones that can lead to serious medical conditions or even death. It is believed that symptoms may appear in 2 to 14 days, as the incubation period for the novel corona virus has not yet been confirmed. However, in India 14 days minimum quarantine period is declared by Government for suspected cases. Since it is a new type of virus, there is a lot of research being carried out across the world to understand the nature of the virus, origins of its spreads to humans, the structure of it, possible cure / vaccine to treat COVID-19. India also became a part of these research efforts after the first two confirmed cases were reported here on January 31, 2020. Then in India screening of traveler at airport migrant was started, immediate Chinese visas was canceled, and who was found affected from COVID-19 kept in quarantine centers (Ministry of Home Affaires Government of India, Advisory).

In continuation, we take a look at few of the interesting and important research being carried out in India with respect to COVID-19. ICMR, India claims that SARI patients with no record of international travel or contact with infected persons tested positive for COVID-19. Hence it is important to optimize testing by developing strategies to identify potential cases that have a higher chance of being infected. Since the availability of the resources like testing kits, labs, health personnel etc. is limited in India as for as concerned the population, the most practical approach is to test symptomatic patients presenting to hospitals, hotspots and aggressive testing to identify and contain local chains of transmission.

In absence of a definite treatment modality like vaccine, physical distancing has been accepted globally as the most efficient strategy for reducing the severity of disease and gaining control over it (Ferguson, 2020; Singh et al., 2020). Also in India it is reported that the country is well short of the WHO’s recommendations of minimum threshold of 2.28 skilled health professionals per 1000 population (Anand et al., 2016). Therefore, on 24 March 2020, the Government of India under Prime Minister Narendra Modi Ji ordered a nationwide lockdown for 21 days, limiting movement of the entire 1.3 billion population of India as a preventive measure against the COVID-19 pandemic in India. It was ordered after a 14-hour voluntary public curfew on 22 March. The lockdown was placed when the number of confirmed positive corona virus cases in India was approximately 500. On 14 April, Prime Minister of India extended the nationwide lockdown until 3 May, with a conditional relaxation after 20 April for some regions. On 1 May, the Government of India again extended the nationwide lockdown further by two weeks until 17 May. Also, the Government has divided the entire nation into three zones viz. green, red and orange with relaxations applied accordingly.

There are already various measures such as social distancing, lockdown masking and washing hand regularly has been implemented to prevent the spread of COVI-19, but in absence of particular medicine and vaccine it is very important to predict how the infection is likely to develop amongst the population that support prevention of the disease and aid in the preparation of healthcare service. This will also be helpful in estimating the health care requirements and sanction a measured allocation of resources. It is well known fact that COVID-19 has spread differently in different countries, any planning for increasing a fresh response has to be adaptable and situation-specific. Data obtained on COVID-19 outbreak have been studied by various researchers using different mathematical models (Rao et al., 2020; Chang et al., 2020). Many other studies (Huang et al., 2020; Hui et al., 2020; Corman et al., 2020; Rothe et al., 2020; Anastassopoulou et al., 2020; Gamero et al., 2020, Singh, 2020) on this recent epidemic have been reported so many meaningful modeling results based on the different principles of mathematics.

Most of pandemics follow an exponential curve during the initial spread and eventually flatten out (Junling et al., 2014). SIR model is one of the best suited models for projecting the spread of infectious diseases like COVID-19 where a person once recovered is not likely to become susceptible to the infection again (Kermack & McKendrick, 1991). Susceptible-Infectious-Recovered (SIR) compartment model (Herbert, 2000) is used to include considerations for susceptible, infectious, and recovered or deceased individuals. These models have shown a significant predictive ability for the growth of COVID-19 in India on a day-to-day basis so far. A recent study by Mandal et al., 2020 has shown that social distancing can reduce cases by up to 62 percent.

Further, time series models have been employed for predicting the incidence of COVID-19 disease. As compared to other prediction models, for instance support vector machine (SVM) and wavelet neural network (WNN), ARIMA model is more capable in the prediction of natural adversities (Zhang et al., 2019) A time dependent SIR models have been defined to observe the undetectable infected persons with COVID-19 (Chen et al., 2020). Chatterjee et al., 2020 studied a stochastic mathematical model of the COVID-19 epidemic in India. The logistic growth regression model is used for the estimation of the final size and its peak time of the corona virus epidemic in many countries of the World and found similar result obtained by SIR model (Batista, 2020).

It is well known that the effects of social distancing become visible only after a few days from the lockdown. This is because the symptoms of the COVID-19 normally take some time to come out after getting infected from the COVID-19. The peak infection is reached at the end of June 2020 with in excess of 150 million infective in India and the total number infected is estimated to be 900 million (Singh et al., 2020). Other estimates indicates that, with hard lockdown and continued social distancing, the peak total infections in India will be 97 million and the number of infective by September is likely to be over 1100 million (Schueller et al., 2020).

Due to the recent development of this pandemic, we are interested in addressing the following important issues about COVID-19:

1. What is the expected time to stop new corona cases?
2. What is the expected maximum number of corona cases?
3. The significance of lockdown.

In this paper, instead of developing a mathematical model for the pattern of spread of COVID-19, an attempt has been made to resolve these above issues in India.

## Data and Methodology

Let us define a function called tempo of disease that is the first differences in natural logarithms of the cumulative corona positive cases on a day, which is as:

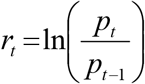

Where *p_t_* and *p_t−1_* are the number of cumulative corona positive cases for period *t* and *t*−1, respectively. When *p_t_* and *p_t−1_* are equal then *r_t_* will become zero. If this value of *r_t_* i.e. zero will continue a week then we can assume no new corona cases will appear further. In the initial face of the disease spread the tempo of disease increases but after sometime when some preventive majors is being taken then it decreases.

Since *r_t_* is a function of time then the first differential is defined as

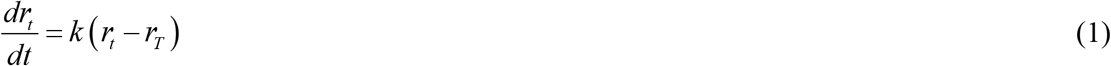

Where *r_t_* denotes the tempo that is the first differences in natural logarithms of the cumulative corona positive cases on a day, *r_T_* is the desired level of tempo i.e. zero in this study, *t* denotes the time and *k* is a constant of proportionality.

Equation 1 is an example of an ordinary differential equation that can be solved by the method of separating variables. The equation 1 can be written as

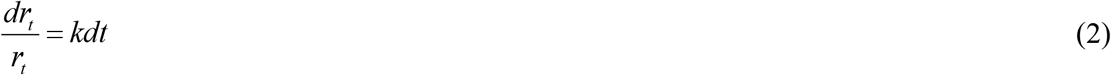

Integrating equation 2, we get

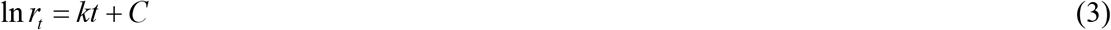

Where *C* is an arbitrary constant. Taking the antilogarithms of both sides of equation 3 we have

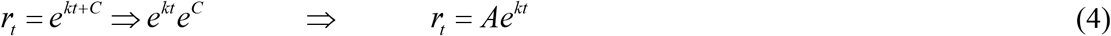

Where *A*=*e^C^*. This equation 4 is the general solution of equation 1. If *k* is less than zero, equation 4 tells us how the corona positive cases will decreases over the time until it reaches zero. Value of *A* and *k* is estimated by least square estimation procedure using the data sets.

The paper used series of daily cases from the website corona19india.org. In this study the day wise cumulative number of corona positive cases from April 1 to May 10, 2020 has been used to know when the tempo of disease will become zero and what will be the size at that time. Also an attempt has been done to understand the significance of lockdown with the help of variation in tempo of disease during the various lockdown periods. The Government of India implemented lockdown on 24^th^ March, 2020 and expected that the tempo of disease is decreasing. We have analyzed India data along with some states such as Uttar Pradesh, Madhya Pradesh, Rajasthan, Bihar, Maharashtra, Gujarat, Delhi, Punjab, West Bengal, Tamil Nadu, Telangana, Karnataka, Andhra Pradesh and Kerala.

## Findings

**Table 1** reveals that some indicators related to the COVI-19 in India and some selected states. The fatality rate is highest in West Bengal followed by Gujarat and Madhya Pradesh. Maharashtra has fatality rate as 3.8 and Uttar Pradesh as 2.1 however in Tamil Nadu and Telangana the fatality rate is 0.7 and 2.6 respectively. Kerala shows 0.8 and Andhra Pradesh 2.3 as Fatality rate. Rajasthan and Delhi has 2.8 and 1.0 the fatality rate. Recovery rate is highest in Kerala followed by Telangana. The lowest recovery rate is found in Maharashtra and West Bengal. In India as whole 30 percent recovery rate has been reported however the fatality rate as 3.3 percent. The reproduction rate of the COVID-19 in India is 1.13 and for states it is maximum (1.15) for Madhya Pradesh and Tamil Nadu. It is lowest For Kerala and Telangana. Large variation observed in the percent confirmed cases among total testing cases, from 1.2 in Bihar to 9.1 in Maharashtra. In Punjab the recovery rate is very low however in Karnataka the percent confirmed among total test is very low means either the disease prevalence is low or the quality of testing kit is not good. In Gujarat and Delhi it is 7.4 and 7.9 percent respectively. These percentages are very low because the testing are done in the hotspots only if the population of hotspot only is considered that these percentages might be more.

**Table 1:** Some indicators related to COVID-19 in India and some states.

| Name of State | Fatality Rate<br>(in percent)<sup>1</sup> | Recovered Rate<br>(in percent)<sup>2</sup> | Percent Confirmed<br>among Total Tests<sup>3</sup> | Reproduction<br>Number<sup>4</sup> |
| --- | --- | --- | --- | --- |
| Telangana | 2.6 | 64.2 | 5.8 | 1.09 |
| Andhra Pradesh | 2.3 | 46.0 | 1.2 | 1.14 |
| Bihar | 0.8 | 54.0 | 1.9 | 1.13 |
| Delhi | 1.0 | 30.9 | 7.9 | 1.13 |
| Gujarat | 6.1 | 26.8 | 7.4 | 1.14 |
| Kerala | 0.8 | 95.8 | 1.4 | 1.05 |
| Madhya Pradesh | 5.8 | 39.0 | 5.4 | 1.15 |
| Maharashtra | 3.8 | 18.8 | 9.1 | 1.13 |
| Rajasthan | 2.8 | 55.4 | 2.4 | 1.11 |
| Tamil Nadu | 0.7 | 27.9 | 2.8 | 1.15 |
| Uttar Pradesh | 2.1 | 43.2 | 2.7 | 1.11 |
| West Bengal | 9.6 | 20.8 | 5.0 | 1.12 |
| Punjab | 1.8 | 8.9 | 4.6 | 1.09 |
| Karnataka | 3.8 | 48.6 | 0.8 | 1.08 |
| **India** | **3.3** | **30.2** | **4.1** | **1.13** |

<sup>1</sup>Fatality Rate = (Total Deaths/Total Cases)\*100<sup>2</sup>Recovery Rate = (Total Recovered/Total Cases)\*100<sup>3</sup>Percent of Confirmed cases among Total Tests = (Confirmed cases/Total Tests)\* 100<sup>4</sup>Reproduction Number = Indicator of current Transmission Rate**Table 2: Expected time to stop new corona cases and maximum number of corona cases**

**Table 2** gives the estimates of time to stop new corona cases and maximum number of corona case in selected states of India. It has been observed that in Kerala and Telangana, there will be no new case of corona virus by end of May and mid June with expected maximum cases 600 and 2000 respectively. Whereas in Uttar Pradesh, Madhya Pradesh, Rajasthan, Karnataka, Punjab, West Bengal, Bihar and Andhra Pradesh, there will be no new case of corona virus by the end July with expected maximum cases 11000, 18000, 9000, 3000, 4000, 16000, 4000 and 4000 respectively. In Delhi, the virus will continue till the mid August. In Maharashtra, Gujarat and Tamil Nadu the pandemic will continue till the end of August with expected number of cases 115000, 45000 and 35000 respectively. It is expected that the COVID-19 virus will disappear in India more or less by the end of August with the maximum number of cases about 350000.

**Table 2:** Expected time to stop new corona cases and maximum number of corona cases.

| State | Expected Time<br>When $r_t = 0$ | Expected Maximum<br>Number of Corona Cases |
| --- | --- | --- |
| Telangana | Mid June | 2000 |
| Andhra Pradesh | July End | 4000 |
| Bihar | July End | 4000 |
| Delhi | Mid August | 33000 |
| Gujarat | August End | 45000 |
| Kerala | May End | 600 |
| Madhya Pradesh | July End | 18000 |
| Maharashtra | August End | 115000 |
| Rajasthan | July End | 9000 |
| Tamil Nadu | August End | 35000 |
| Uttar Pradesh | July End | 11000 |
| West Bengal | July End | 16000 |
| Punjab | July End | 4000 |
| Karnataka | July End | 3000 |
| **India** | **August End** | **350000** |

Government suggested and implemented social distancing and lockdown to control the spread of COVID-19 in the society. In **Table 3** an attempt has been made to show the summary statistics of tempo of COVID-19 *r_t_* during various lockdown periods in India. It is observed that average tempo is maximum (0.167 with standard deviation 0.062) in the period prior to the lockdown. During the first lockdown period the average tempo is 0.140 with standard deviation 0.044 however in lockdown 2 it is 0.070 with standard deviation 0.012, thus it is clear that both average and standard deviation are decreasing. **Table 4** represents the results of ANOVA testing for mean of *r_t_* during various lockdown periods which is significant means that the average tempo of COVID-19 is significantly different is various periods considered. A group wise comparison of the average tempo of COVID-19 *r_t_* during various lockdown periods is shown in **Table 5** which reveal that lockdown is significantly affects the spread of COOVID-19. **Figure 1** show that tempo of disease *r_t_* is declining towards zero with time, more rapidly in Kerala and Telangana than other states. Whereas, in rest of the states it is declining slowly towards *r_t_* = 0.

**Table 3:** Summary of *r_t_* during various lockdown period.

| Lockdown Period | N | Mean | Std. Deviation | Std. Error | 95% Confidence Interval for Mean |  |
| --- | --- | --- | --- | --- | --- | --- |
|  |  |  |  |  | Lower Bound | Upper Bound |
| No | 11 | 0.167 | 0.062 | 0.019 | 0.125 | 0.209 |
| 1 | 22 | 0.140 | 0.044 | 0.009 | 0.120 | 0.159 |
| 2 | 19 | 0.070 | 0.012 | 0.003 | 0.064 | 0.076 |
| Total | 52 | 0.120 | 0.057 | 0.008 | 0.104 | 0.136 |

**Table 4:** ANOVA test for mean of *r_t_* during various lockdown period.

| Source of Variations | Sum of Squares | df | Mean Square | F | p value |
| --- | --- | --- | --- | --- | --- |
| Between Groups | 0.081 | 2 | 0.041 | 240.152 | 0.000 |
| Within Groups | 0.082 | 49 | 0.002 |  |  |
| Total | 0.163 | 51 |  |  |  |

**Table 5:** Group wise comparison of means of *r_t_* during various lockdown periods.

| (I) group | (J) group | Mean Difference (I-J) | Std. Error | p value | 95% Confidence Interval |  |
| --- | --- | --- | --- | --- | --- | --- |
|  |  |  |  |  | Lower Bound | Upper Bound |
| No | 1 | 0.027 | 0.015 | 0.237 | -0.010 | 0.065 |
|  | 2 | 0.097 | 0.016 | 0.000 | 0.059 | 0.136 |
| 1 | 2 | 0.070 | 0.013 | 0.000 | 0.038 | 0.102 |

**Figure 1:**
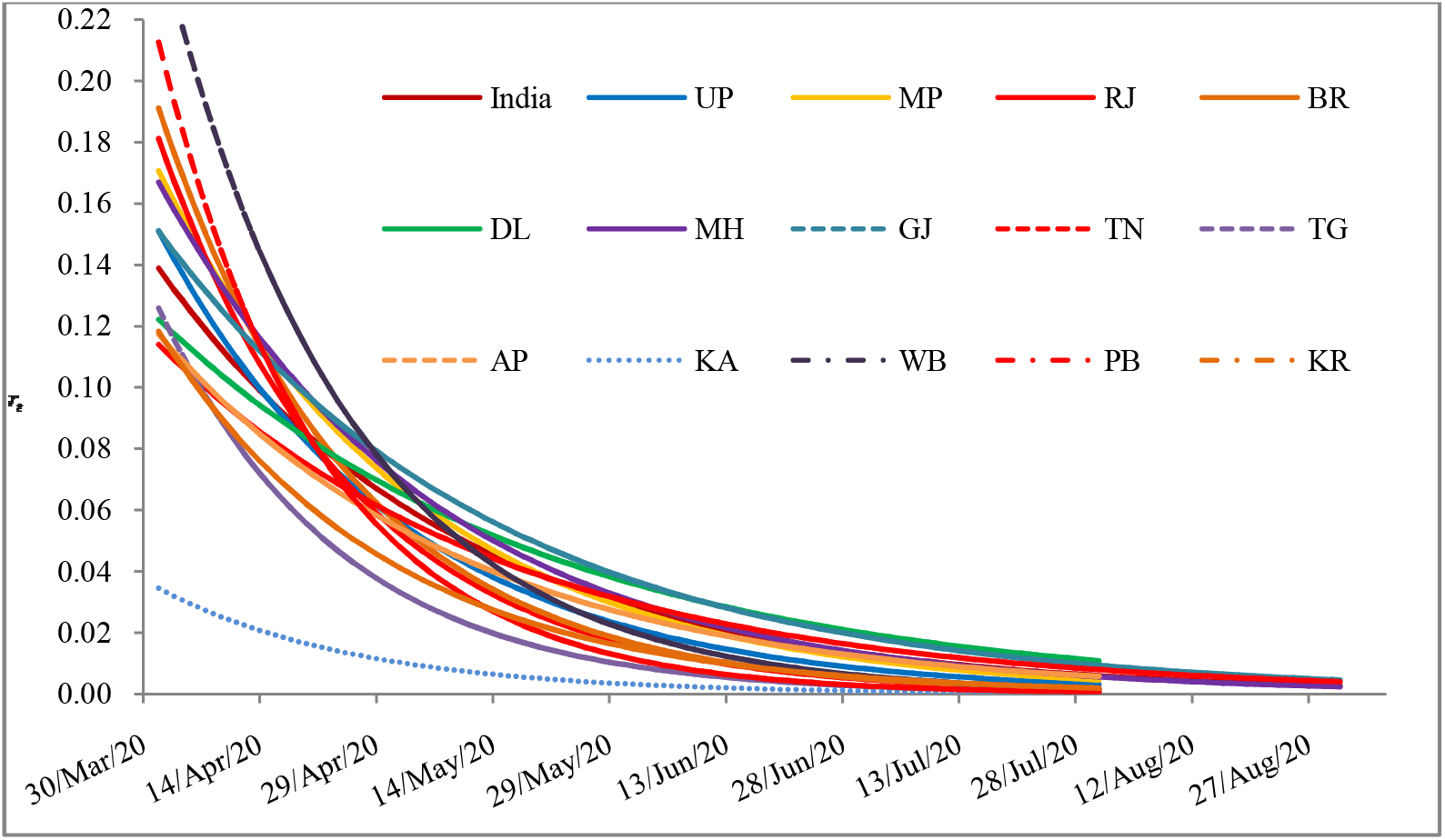
Trend of *r_t_* for India and some states

## Conclusion

COVID-19 has been declared as pandemic by WHO and is currently become a major global threat. Prediction of a disease may help us to understand the factors affecting it and the steps that we can take to control it. The Government of India has taken preventive measures such as complete lockdown in the very early stage of disease, physical distancing and case isolation. The most important issue is that many healthcare professionals are visiting each and every household in the hotspot area across the country to trace and isolate infected persons to curtail the spread of disease. In order to support the prevention of the disease and aid to the healthcare professionals, an attempt has been made to develop a simple model for the prediction of confirmed COVID-19 cases and to utilize that model for forecasting future COVID-19 cases in India. As per the model forecast, the confirmed cases are expected to gradually decrease in the coming weeks. It is also likely that the efforts such as lockdown and physical distancing affect this prediction start to decline. On the basis of considered data, one can predict that the final size of corona virus pandemic in India will be around 350000 by the end of August. The exponential model used in this study is a data driven model. Thus, its forecasts are as reliable and can capture the dynamics of the pandemic. Due to real time change in data daily, the predictions will accordingly change. Hence, the results from this paper should be used only for qualitative understanding.

## Data Availability

Data is available on line covid19india.org

